# Diagnostic accuracy of MRI compared to CTPA for pulmonary Embolism : A meta analysis

**DOI:** 10.1101/2023.08.02.23293299

**Authors:** Dev H. Desai, Abhijay Shah, Hetvi Shah, Aarya A. Naik, Sharif Mohammed Sadat, Dwija Raval

## Abstract

**Background:** This meta-analysis presents a comparison between Computed Tomography Pulmonary Angiography (CTPA) and Magnetic Resonance Angiography (MRA), to diagnose a pulmonary embolism. Computed tomography presents the advantage of imaging the entire thorax, facilitating the diagnosis of conditions that are commonly mistaken for pulmonary embolism, such as pneumonia, aortic dissection, and malignancy. UK and US guidelines have established CT amongst the basic investigations for pulmonary embolism. MRA does not require the use of ionizing radiation or iodinated contrast, thus making it possible for routine use of multiphasic acquisitions as well as for repeated contrast injections

**Methodology:** For the collection of the data, a search was done by two individuals using PubMed, Google Scholar, and Cochrane Library databases for all relevant literature. Full - Text Articles written only in English were considered. Each qualifying paper was independently evaluated by two reviewers. Each article was analyzed for the number of patients, their age, procedure modality, and incidence of the pre decided complications.

**Results:** The results also showed a high positive predictive value of 0.947 or 94.7% for MRA in the diagnosis of Pulmonary embolism, as compared with CTPA. Some analyses have marked MRA to have low specificity. These results establish MRA as a respectable alternative for diagnosis of APE, especially in cases when reducing radiation exposure is desired. However, the gold standard of diagnosis remains Computed Tomography Pulmonary Angiography.

**Conclusion:** As the results show, though MRA has high statistical value for the diagnosis of pulmonary embolism, it also has its drawbacks. MRA cannot be used in severely ill patients as it continues to be challenging, with the longer scan times and multiple breath holds that are required in different MR protocols are difficult to follow in these patients. CTPA remains the gold standard for diagnosis of Pulmonary embolism, with MRA as a secondary test used when CTPA is contraindicated.

## INTRODUCTION

Pulmonary embolism covers a wide spectrum of presentations, ranging from asymptomatic individuals to life-threatening medical emergencies

The importance of identifying pulmonary artery embolism (PE) is well established. Unfortunately, symptoms are often not specific, making a diagnosis of PE very challenging, based only on clinical presentation. The common symptoms such as acute chest pain and dyspnea are not specific to PE. As a result, imaging is frequently requested to exclude pulmonary embolus. [1]

Pulmonary embolus (PE) is estimated to cause from 200,000 to 300 000 deaths a year. The patients at risk are generally hemodynamically unstable. The choice of treatment in PE depends on the risk of poor result. Hypotension is the most significant predictor of poor outcome and defines those with massive PE.[2][3]

Overall, the diagnosis of Pulmonary embolism involves a combination of diagnostic tests, including CTPA, MRA, ultrasound, blood tests, and chest X-ray. The choice of test may depend on factors such as the patient’s medical history, the availability of imaging equipment, and the expertise of the medical team Computed tomography presents the advantage of imaging the entire thorax, facilitating the diagnosis of conditions that are commonly mistaken for pulmonary embolism, such as pneumonia, aortic dissection, and malignancy. UK and US guidelines have established CT amongst the basic investigations for pulmonary embolism.[2]

In critically ill patients, computed tomographic pulmonary angiography appears to be the investigation of choice for many physicians. In patients of renal insufficiency or contrast allergy, ventilation-perfusion scans are an alternative choice that may be used.

In patients who are unstable, and cannot be shifted for radiological tests, echocardiography, especially transesophageal echocardiography is an option.

[4] A positive Doppler ultrasound of the lower extremity indicating a deep vein thrombosis in the setting of chest pain and dyspnea may also help in the decision for treatment of a pulmonary embolism

Another alternative to computed tomography angiography is contrast enhanced magnetic resonance angiography (CT-MRA). With the recent enhancements in scanner technology, recent studies have shown great capacity in detecting pulmonary embolisms. [5]

MRA does not require the use of ionizing radiation or iodinated contrast, thus making it possible for routine use of multiphasic acquisitions as well as for repeated contrast injections.[6] Magnetic resonance pulmonary angiography(MRA) and magnetic resonance venography (MRV) when performed together, have a higher sensitivity than magnetic resonance pulmonary angiography alone in patients with technically adequate images. However, it is harder to obtain technically adequate images with the combination of procedures.

## METHODOLOGY

### DATA COLLECTION

For the collection of the data, a search was done by two individuals using PubMed, Google Scholar, and Cochrane Library databases for all relevant literature. Full - Text Articles written only in English were considered.

The medical subject headings (MeSH) and keywords ‘CT Pulmonary Angiogram’, ‘MR Pulmonary Angiography’, and ‘Pulmonary Embolism’were used. References, reviews, and meta-analyses were scanned for additional articles.

### INCLUSION AND EXCLUSION CRITERIA

Titles and abstracts were screened, and Duplicates and citations were removed. References of relevant papers were reviewed for possible additional articles. Papers with detailed patient information and statically supported results were selected.

We searched for papers that show more accurate diagnoses, where procedures considered were CTPA and MRPA.

The inclusion criteria were as follows: (1) studies that provided information about the accurate diagnosis with CTPA and MRPA; (2) studies published in English; (3) Studies comparing MRPA with CTPA as a Diagnosis modality for cases of pulmonary embolism.

The exclusion criteria were: (1) articles that were not full text, (2) unpublished articles, and (3) articles in other languages.

### DATA EXTRACTION

Each qualifying paper was independently evaluated by two reviewers. Each article was analyzed for the number of patients, their age, procedure modality, and incidence of the pre decided complications. Further discussion or consultation with the author and a third party was used to resolve conflicts. The study’s quality was assessed using the modified Jadad score. In the end, According to PRISMA, a total of 39 RCTs with a total of 6273 patients were selected for further analysis.

### ASSESSMENT OF STUDY QUALITY

Using the QualSyst tool, two writers independently assessed the caliber of each included study. This test consists of 10 questions, each with a score between 0 and 2, with 20 being the maximum possible overall score. Two authors rated each article independently based on the above criteria. The interobserver agreement for study selection was determined using the weighted Cohen’s kappa (K) coefficient. For deciding the bias risk for RCTs, we also employed the Cochrane tool. No assumptions were made about any missing or unclear information. there was no funding involved in collecting or reviewing data.

### STATISTICAL ANALYSIS

The statistical software packages RevMan (Review Manager, version 5.3), SPSS (Statistical Package for the Social Sciences, version 20), Google Sheets, and Excel in Stata 14 were used to perform the statistical analyses. The data was obtained and entered into analytic software[7] [21]. Heterogeneity and the diagnostic accuracy were performed. the latter of which was calculated by pooled estimates of sensitivity, specificity, positive predictive value (PPV), diagnostic odds ratios (DOR), and relative risk (RR) with 95 percent confidence intervals to examine critical clinical outcomes (CIs). Diagnosis accuracy and younden index were calculated for each result. Individual study sensitivity and specificity were plotted on Forest plots and in the receiver operating characteristic (ROC) curve. The forest plot and Fagan’s Nomogram were used to illustrate the sensitivity and specificity of different papers.

Heterogeneity was detected by Cochrane Q test and I2 (inconsistency) statistics, with PL<L.10 or I2L>L50%, indicating a significance in heterogeneity.[10] Furthermore, if there was significant heterogeneity (I2L>L50% or PL≤L.05), the random-effects model (DerSimonian-Laird method) was preferred over the fixed-effects model (Mantel-Haenszel method); otherwise, the fixed-effects model was the first choice.

### BIAS STUDY

The risk of bias was evaluated by using QUADAS-2 analysis. This tool includes 4 domains as Patient selection, Index test, Reference standard, Flow of the patients, and Timing of the Index tests.

## RESULT

### MRPA v CTPA

Here, Table 1 describes all the descriptions of papers used for the MRPA vs CTPA study. All the results described above, in the forest chart (figure 2), the comparison of the sensitivity and specificity of different papers can be observed. The same is illustrated in the SROC curve. (Figure 3). A total of 39 RCTs with 6273 were selected for the study, out of which 14 studies showed sensitivity above 95%, and 23 studies showed specificity above 95%. There were 9 studies that showed both sensitivity and specificity, to be over 95%. The value of True Positive (TP) was 2172, that of True Negative (TN) was 3697, that of False Positive (FP) was 121, and that of False Negative (FN) was 283. With a confidence interval 95%, sensitivity, specificity and positive predictive values were calculated, a summary of which is available in figure 2. The sensitivity of MRPA is 0.885, with a CI of 95% in a range of 0.834 to 0.936; the mean being 0.051. The specificity of MRPA is 0.968, with a CI of 95% in a range of 0.927 to 1.010; the mean being 0.042.

**Table 1.**
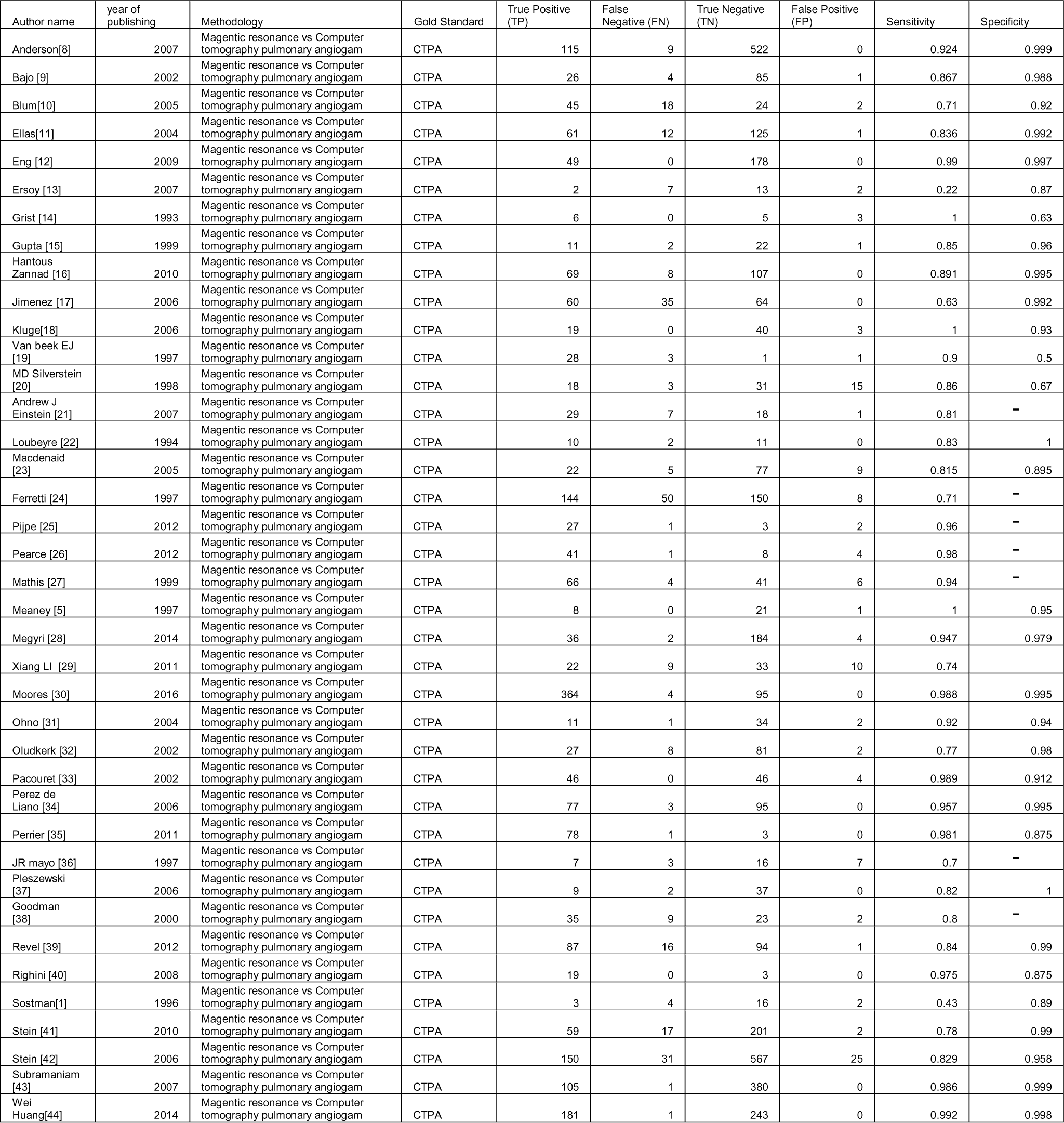
Table of description.

**Figure-1.**
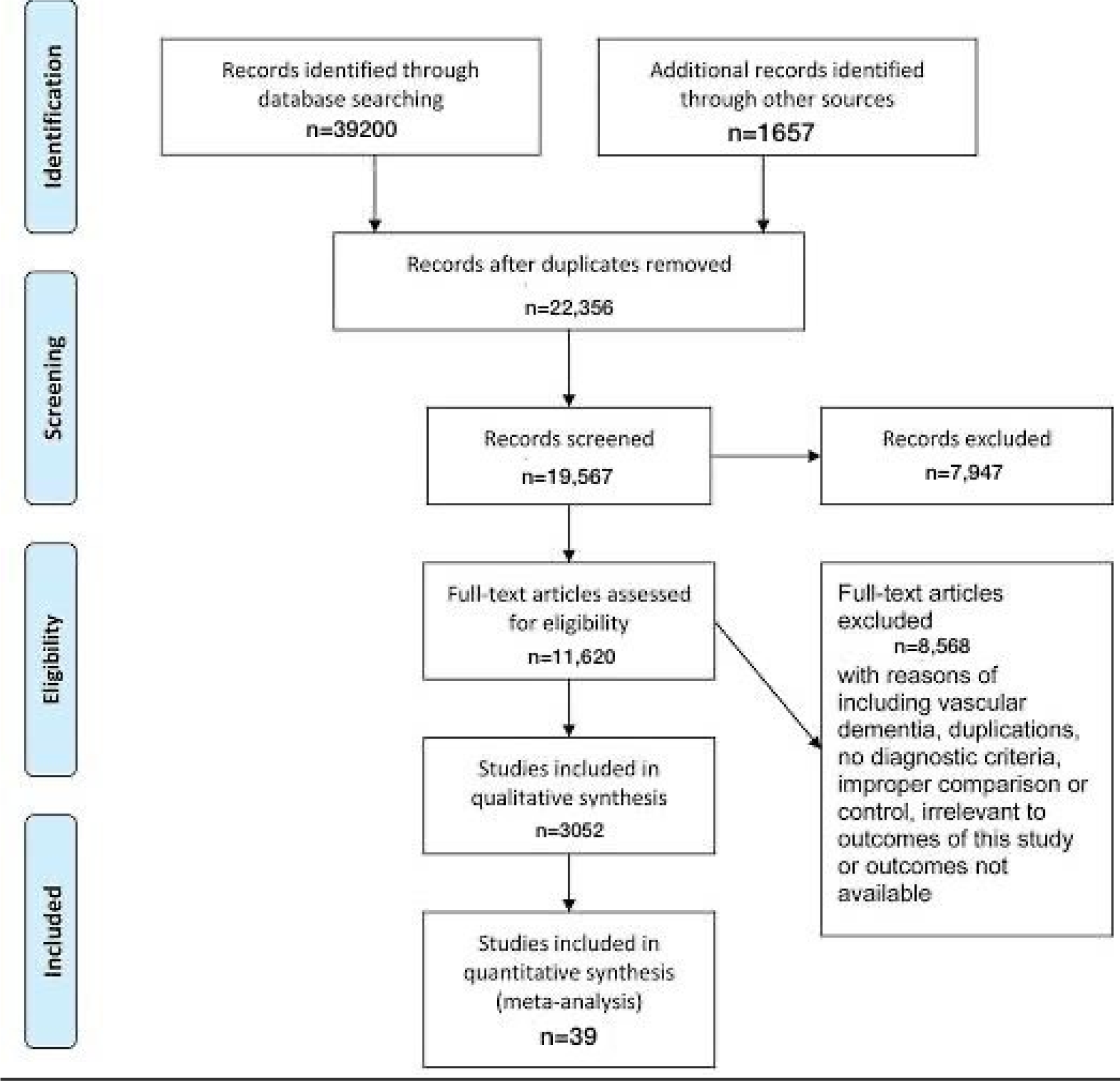
Prisma chart.

**Figure 2:**
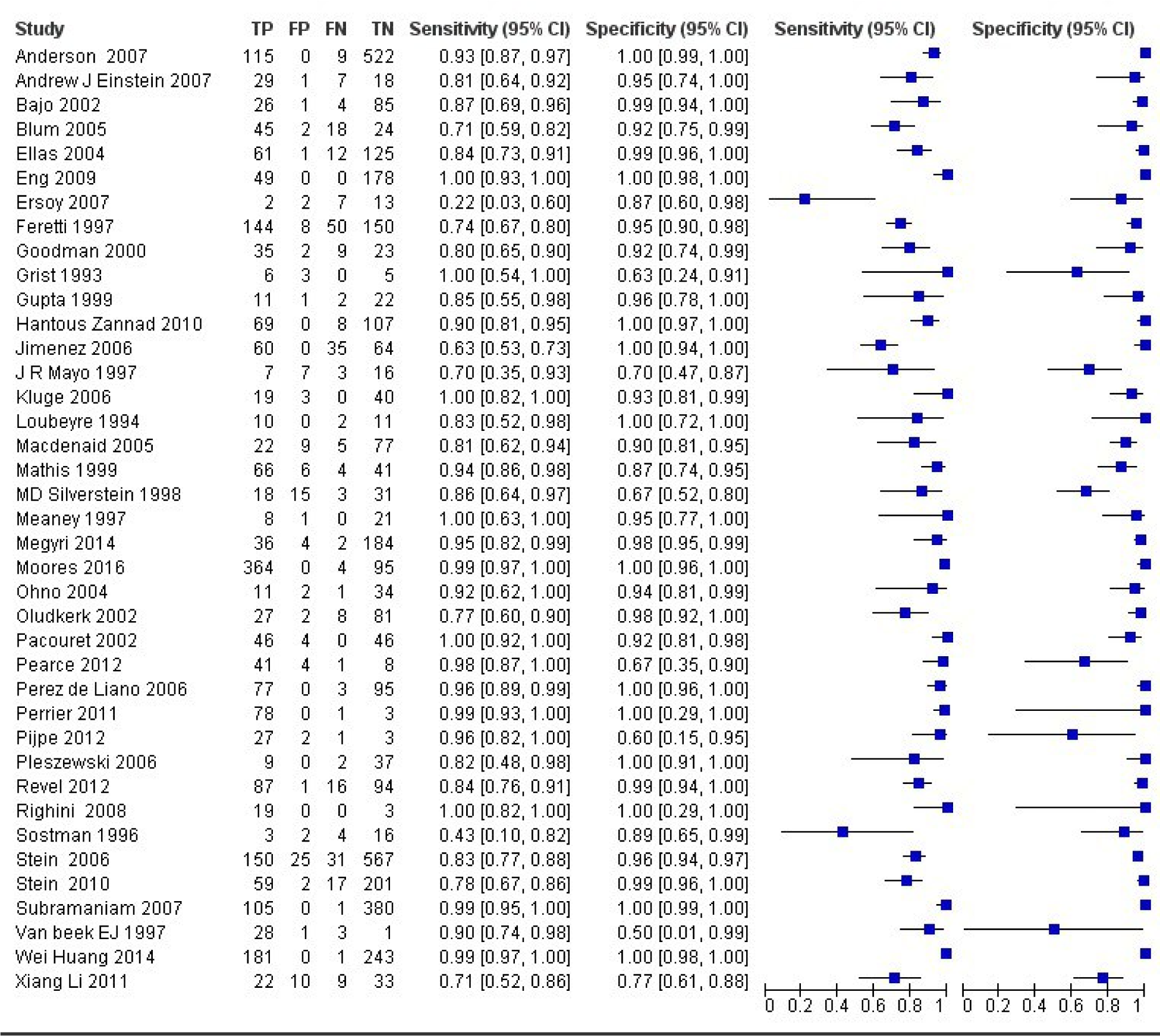
The forest chart summary for MRPA and CTPA.

**Figure 3:**
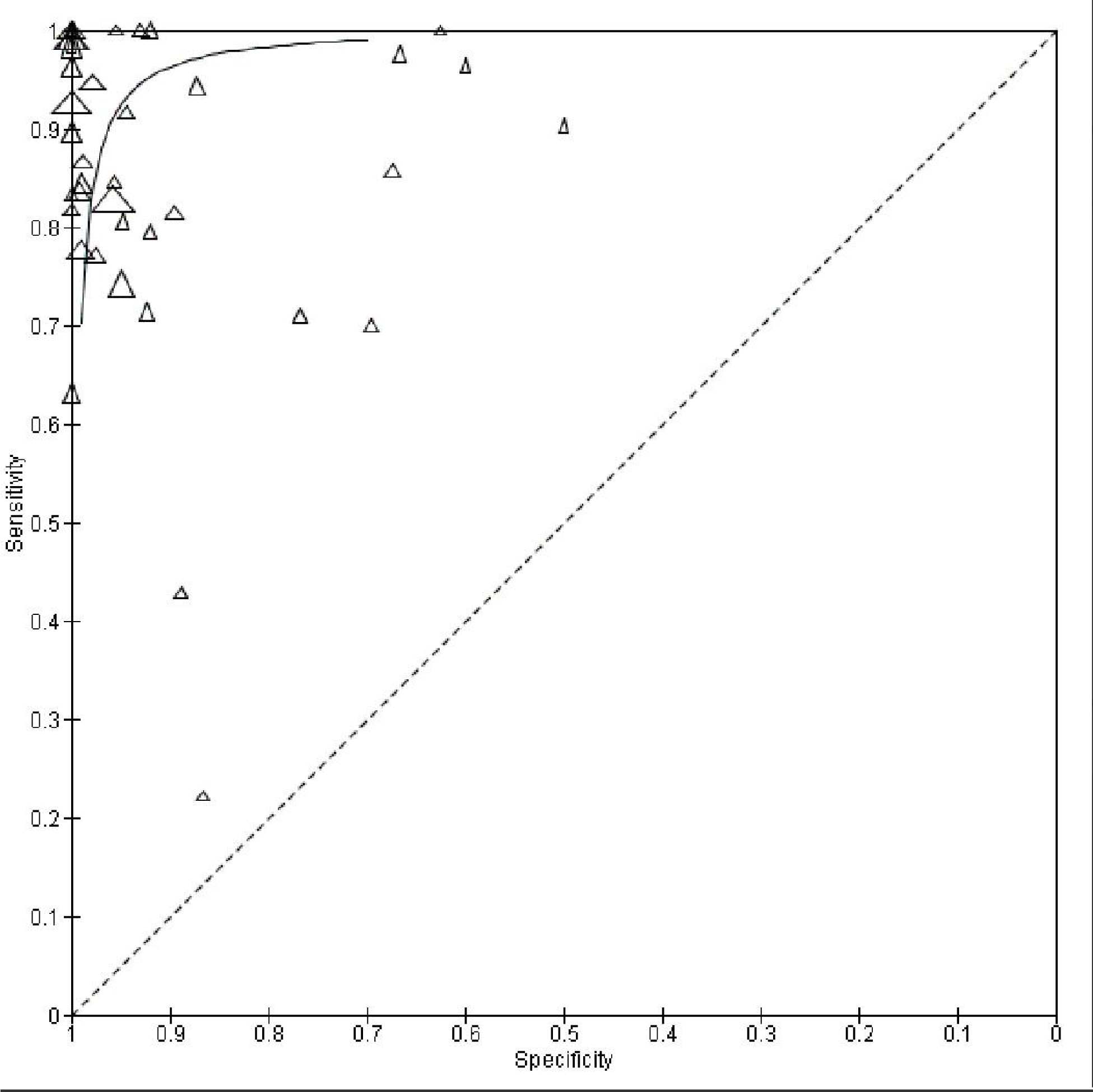
The SROC plot summary for MRPA vs CTPA.

The summary of the ROC curve is described in figure 3. It shows that the area under the curve for MRPA was 0.9265 and the overall diagnostic odds ratio (DOR) was 142.67 (CI95 = 76.10 to 267.48) with index Q* of intersection at 0.9128 (SE=0.0159) and Younden Index being 0.853

P value for Cochrane Q for DOR was lower than 0.0001 and I^2^ was at 76.3%

Figure 4 describes the summary of Fagan plot analysis for all the studies considered for MRPA vs CTPA, showing a prior probability of 39% (0.6); a Positive Likelihood Ratio of 28; a probability of post-test 95% (18.0); a Negative likelihood ratio of 0.12, and a probability of post-test 7%(0.1).

**Figure 4:**
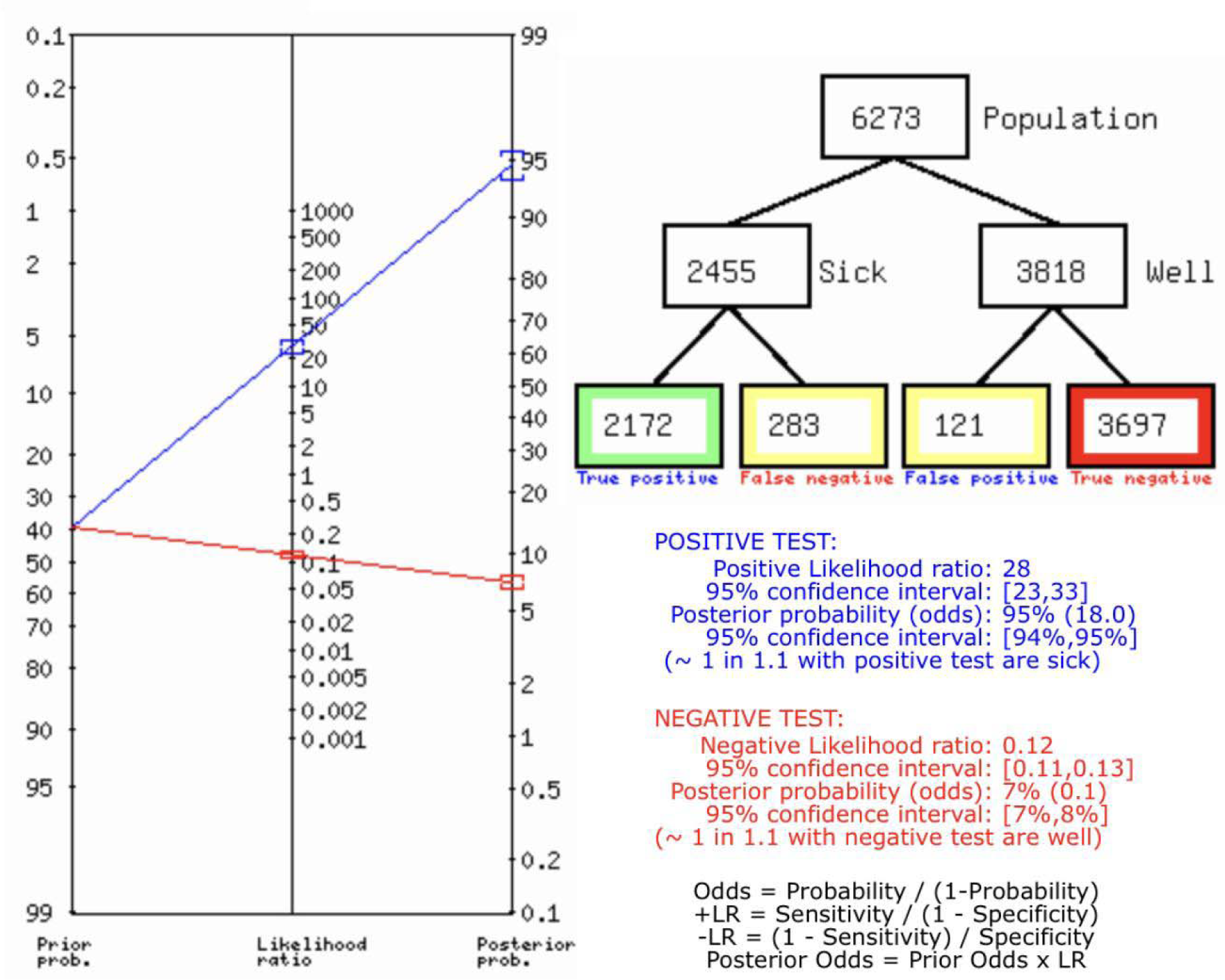
Fagan’s Analysis for MRPA vs CTPA.

Figure 5 describes the bias study and applicability concern. The tool used for this is QUADAS-2 analysis. The study is having low risk of bias and low risk of applicability concern. The reference standard is suggestive of risk of bias and having low applicability concern.

**Figure 5.**
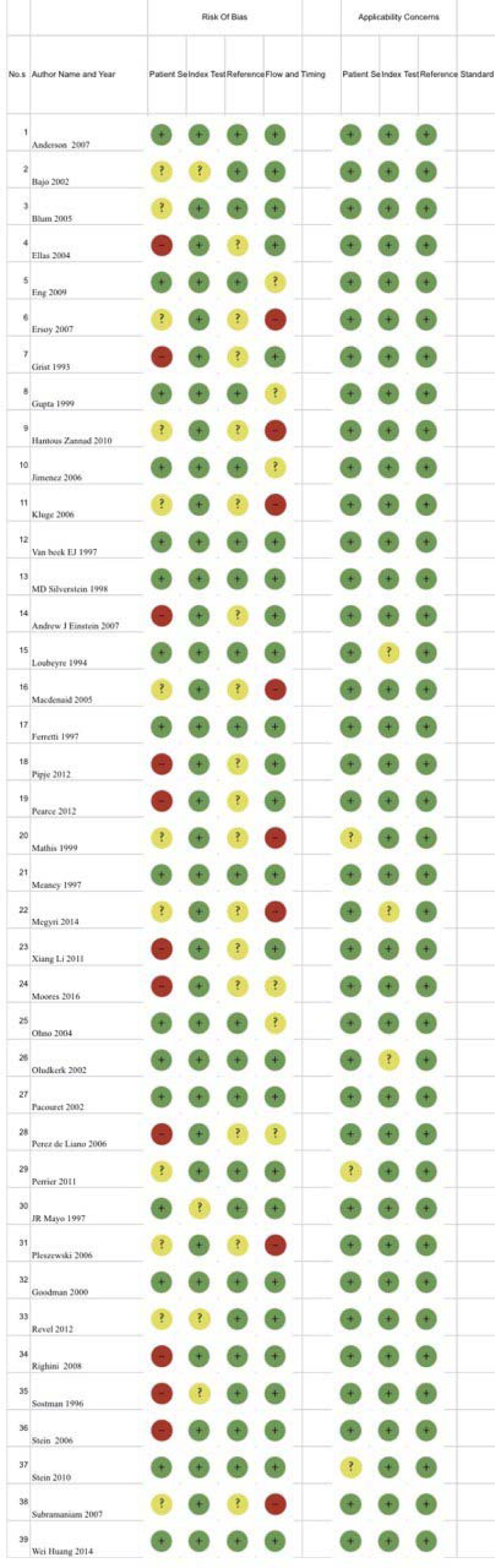
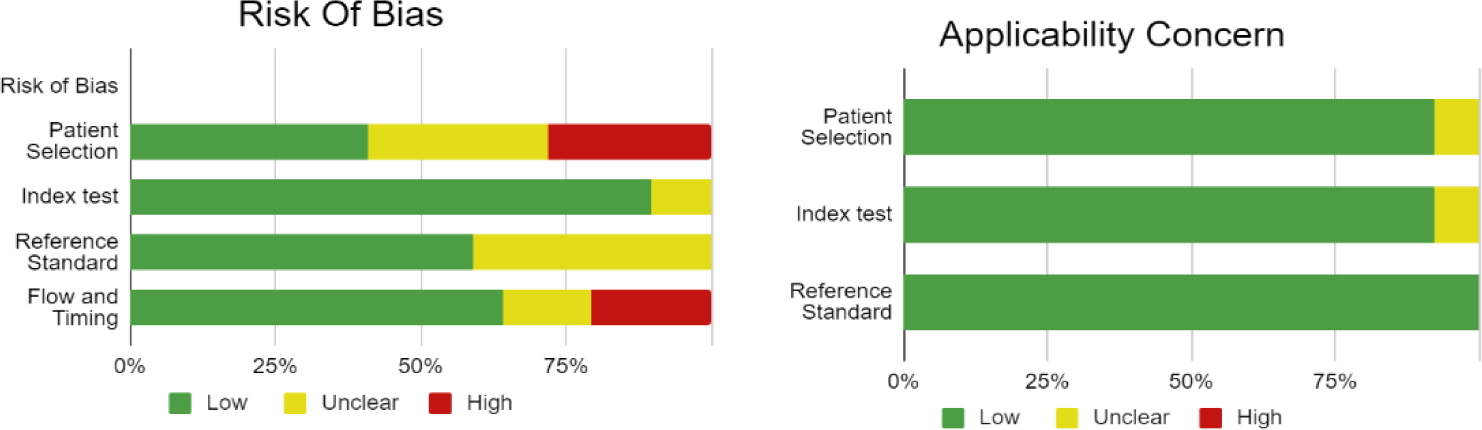
Risk of Bias and Applicability concern.

## DISCUSSION

Pulmonary embolism is a sudden and potentially life-threatening condition that occurs due to blockage of one or more pulmonary arteries from embolic material. The embolus or blood clot generally forms in one of the deep veins of the legs or pelvis. This blockage reduces or stops blood flow and can elevate pressures within the right ventricle of the heart.

It is an often underdiagnosed condition, due to its non specific signs and symptoms. Thus, though early treatment is very effective, diagnosis using objective tests that can establish or refute diagnosis are the standard of care. Diagnosis should be based on the clinical probability of PE.[6]

This meta-analysis presents a comparison between Computed Tomography Pulmonary Angiography (CTPA) and Magnetic Resonance Angiography (MRA), to diagnose a pulmonary embolism.

This review has many strengths, chief among which is the systematic approach to identifying and including comprehensive studies, which makes it unlikely that relevant studies were not included.

Computed tomography pulmonary angiography has made the diagnosis of pulmonary embolism in clinically suspected cases easier. CTAP has high sensitivity and specificity, which are comparable with invasive pulmonary angiography. It has been shown to confirm or rule out APE with satisfactory certainty, removing the need for additional imaging tests. Nevertheless, inherent limitations of CT include substantial radiation exposure, thus, alternative tests and imaging are still encouraged.[45]

One such test with a high rate of success in diagnosis is MRA.

### 2.1 Our results

This meta-analysis was conducting using 39 scientific papers. After thorough perusal of each scientific paper, the data was collated and statistical tests were conducted.

The established gold standard for diagnosis of pulmonary embolism continues to be Computed Tomography. However, in recent years, the use of MRA has increased, with promising results. This meta-analysis showed a high sensitivity (0.885) as well as high specificity (0.968) for MRA. The results also showed a high positive predictive value of 0.947 or 94.7% for MRA in the diagnosis of Pulmonary embolism, as compared with CTPA.

These results establish MRA as a respectable alternative for diagnosis of APE, especially in cases when reducing radiation exposure is desired.

### 2.2 Compare with other papers

A recent analysis indicated that on a patient-based level, MRI can yield high diagnostic accuracy in the detection of acute pulmonary embolism, especially in images that are technically adequate. MRA carries a more attractive potential as an alternative diagnostic test to CTAP due to the lack of ionizing radiation that is associated with CTAP, as well as lower risk of complications that are linked with use of contrast media.

Inconclusive MRI examinations can occur due to many reasons, including poor arterial opacification or artifact in the plate due to motion at the time of scan.

A combination of MRI tests with greater ability to differentiate, than single techniques appears to be more promising in the diagnosis of PE.

From a vessel based perspective, MRI exhibits better diagnostic capability in for embolisms in proximal arteries, but has low sensitivity for peripheral embolisms.[45][46]CTPA is routinely applied in practice, even in severe cases, emergency settings and dyspneic patients, as it only requires short infrequent breath holding. However, in patients who have contraindications to use of contrast material, such as allergies or younger patients, MRA is an alternative method for pulmonary imaging, as it does not require ionizing radiation or contrast material.

However, the use of MRA in severely ill patients continues to be challenging, as the longer scan times and multiple breath holds that are required in different MR protocols are difficult to follow in these patients. [26]Other analyses have marked MRI to have high specificity but limited or low sensitivity in the diagnosis of PE.

A major limitation for the practical application of MRA is inconclusive results. [33] Thus, as the results show, in diagnosis of Pulmonary Embolism, Magnetic Resonance Angiography is a good alternative, but the gold standard of diagnosis remains Computed Tomography Pulmonary Angiography.

## CONCLUSION

As the results suggest, MRA stands to be a good test for the diagnosis of pulmonary embolism when CTPA cannot be used. MRA has been noted to have lower sensitivity in other analyses, as well as to have inconclusive results. CTPA is routinely applied in practice, even in severe cases, emergency settings and dyspneic patients, as it only requires short infrequent breath holding. CTPA can be used in severely ill patients as well, with its shorter test times. MRA may be used in patients with contraindications to use of contrast material, as well as in patients who require lower radiation exposure. This meta-analysis found a high positive predictive value of 94.7% for MRA. However, CTPA remains the gold standard for the diagnosis of PE with its higher sensitivity and specificity for the diagnosis of PE, as well as the ease of testing for patients and physicians.

## Data Availability

All data produced in the present work are contained in the manuscript

